# Longitudinal Digital Phenotyping of Circadian Rest-Activity Rhythms via Wearables as Biomarkers for Late-Life Function, Cognition, and Neuropsychiatric Health

**DOI:** 10.1101/2025.09.20.25336210

**Authors:** Jinjoo Shim, J.P. Onnela

## Abstract

**Background:** Circadian rest-activity rhythmicity, a manifestation of circadian rhythms, characterizes 24-hour activity patterns. Growing evidence links disruption of circadian rhythms in late life to adverse outcomes, including functional and cognitive declines. Yet, most studies have been cross-sectional or restricted to single monitoring period, lacking longitudinal assessment. Consequently, it remains unknown whether deterioration or improvement in circadian rest-activity rhythmicity reflects late-life vulnerability in aging populations.

**Methods:** We analyzed data from the National Health and Aging Trends Study (NHATS), a nationally representative U.S. cohort of adults aged ≥70 years. In Cycles 11 and 12, participants completed 7-day wearable monitoring and assessments of functional, cognitive, and neuropsychiatric outcomes. Circadian biomarkers were derived using cosinor and non-parametric methods (amplitude, MESOR, acrophase, pseudo-F, RA, IS, IV, M10, L5). Participants were classified into four transition groups (Optimal, Improved, Deteriorated, Adverse). Survey-weighted regression and Cox models adjusted for demographics and comorbidities estimated associations, with multiple testing correction.

**Results:** At baseline, weaker rhythm intensity (low amplitude, MESOR, M10, RA) and poor stability (high fragmentation, low regularity) were associated with greater ADL disability, lower SPPB, weaker grip strength, and poorer recall. Longitudinal analyses revealed a graded hierarchy of risk. Participants with deteriorating rhythms represented the most dynamic risk state: declining amplitude was linked to reduced SPPB (β= –0.71, 95% CI: – 0.98 to –0.44) and diminished immediate recall (β= –0.60, –0.91 to –0.29). Declining regularity was also associated with impaired function (SPPB: β= –0.52, –0.82 to –0.21) and cognition (immediate recall: β= –0.43, –0.68 to –0.18; delayed recall: β= –0.45, –0.70 to – 0.20). By contrast, rhythmicity improvement aligned with stabilization. For neuropsychiatric outcomes, high fragmentation was linked to probable dementia, while poor regularity was tied to anxiety/depression.

**Conclusion:** Circadian rest-activity rhythmicity is a robust and dynamic determinant of late-life outcomes. Persistent weakness or deterioration in intensity and regularity was linked to accelerated decline in function and cognition, whereas improvement aligned with stabilization. These findings position circadian rhythmicity as both an early biomarker of vulnerability and a potential modifiable target. Digital phenotyping via wearables offers a scalable, noninvasive framework for early risk detection, personalized intervention, and resilience promotion in aging populations.

## BACKGROUND

Circadian rhythms are fundamental regulators of human physiology and behavior, governing sleep-wake cycles, rest-activity patterns, metabolism, immune function, cognition, and neuropsychiatric health (1–4). These approximately 24-hour rhythms are orchestrated by the central pacemaker within the suprachiasmatic nucleus of the hypothalamus, which synchronizes biological and behavioral processes to environmental zeitgebers, most prominently the light-dark cycle (5). The integrity of circadian organization is essential for maintaining physiological homeostasis; however, with advancing age, rhythms progressively weaken, exhibit increased fragmentation, and lose amplitude and robustness (6,7). Compounding this biological vulnerability, modern lifestyle such as artificial lighting, shift work, and jet lag further obscure temporal boundaries between day and night, accelerating the deterioration of circadian rhythmicity (8–10).

A growing body of evidence highlights the broad health consequences of disruption in late life. Studies have shown that disrupted circadian rest-activity rhythmicity in older adults is linked to functional decline, impaired cognition, frailty, and increased susceptibility to disease (11–15). This existence of this vulnerability is further supported by a large genome-wide association study of 71,500 middle-aged and elderly individuals that revealed associations between genetic risk of circadian disruption and mood disorders (16). Together, these findings indicate that circadian dysfunction is not only correlated with adverse outcomes but also reflects underlying systemic vulnerability. Consequently, circadian health emerges as a central determinant of aging trajectories in late life, with potential utility both as a measurable physiological marker of health deterioration and as an early-warning signal preceding the onset of age-related diseases.

Traditional assessment of circadian rhythms has relied on biomarkers such as melatonin secretion or core body temperature. While biologically precise, these methods are invasive, resource-intensive, and impractical for longitudinal, population-based monitoring. In contrast, wearable technologies, ranging from research-grade actigraphy to increasingly prevalent consumer-grade smartwatches, enable high-resolution, continuous, and noninvasive monitoring of circadian rest-activity rhythmicity through digital phenotyping (17,18). Defined as “the moment-by-moment quantification of the individual-level human phenotype in situ using data from personal digital devices”, digital phenotyping provides a scalable, low-burden approach to capture the dynamics in large populations (19,20). This methodology is particularly well suited for elderly cohorts, as it allows precise and objective characterization of circadian rest-activity rhythmicity under real-world conditions while minimizing participant burden (21,22).

Yet, despite these advances in measurement, most prior studies have been limited to a single seven-day period of wearable activity monitoring, analyzed largely cross-sectionally. Such study designs provide only a static snapshot of circadian rest-activity rhythms and fail to capture within-person changes over time. This is a critical limitation in late life, where dynamic deterioration of circadian organization may more tightly signal impending functional and cognitive decline (12,23). Furthermore, circadian disruption in older adults exerts cumulative effects across the brain, endocrine, immune, and musculoskeletal systems, amplifying its impact throughout the aging process (24). These gaps highlight the need for repeated measurements to establish the relationship between circadian disruption and subsequent functional and cognitive decline, a link that remains largely unknown.

To address this gap, we leveraged longitudinal data from the National Health and Aging Trends Study (NHATS), which includes repeated assessments of circadian activity and health outcomes in adults aged 70 years and older. Participants completed annual research-grade actigraphy monitoring along with detailed assessments of functional capacity and cognitive performance during two consecutive NHATS waves - Cycles 11 and 12. This design enables us to examine both baseline circadian rest-activity rhythmicity and within-person trajectories over time, and to evaluate whether circadian deterioration is longitudinally associated with functional and cognitive decline. We hypothesize that older adults with disrupted rhythms would experience greater deterioration in functional and cognitive outcomes, whereas improvements in rhythmicity would mitigate the risk of age-related deterioration.

## METHODS

### Data source and study participants

We leveraged data from the National Health and Aging Trends Study (NHATS), a prospective, nationally representative cohort of community-dwelling older adults aged ≥70 years in the United States, initiated in 2011. NHATS employs a complex sampling design stratified by age group, race/ethnicity, and geographic region to ensure representativeness of the U.S. older adult population (25,26). Annual interviews are conducted with participants or proxies, collecting information on functional, cognitive, and health outcomes (25). NHATS was approved by the Johns Hopkins Bloomberg School of Public Health Institutional Review Board (IRB No. 00002083) (25). Participants in NHATS completed an informed consent. All methods were performed in accordance with the Declaration of Helsinki (27).

In Cycles 11 (2021) and 12 (2022), a subset of participants was invited to participate in wearable monitoring (28). During the in-home interview, participants provided informed consent and were fitted during their in-home interview with a triaxial, water-resistant, research-grade wrist accelerometer (Actigraph CentrePoint Insight Watch or “Activity Watch”) on their non-dominant wrist for the subsequent seven days (eight total days including the interview day) (28). They were instructed to wear the device continuously (24 hours/day), with removal permitted only for swimming or bathing lasting longer than 30 minutes (28). The Activity Watch passively and continuously recorded participants’ movement in units of gravity (g) at a sampling rate of 64 Hz (28).

Of 1,000 participants selected in Cycle 11, 944 completed interviews and 872 were eligible to wear the accelerometer. Among these, 747 participants (86%) successfully completed the monitoring and returned their device. In Cycle 12, of the 747 participants, 696 were eligible (24 deceased, 27 were ineligible for interview), and 639 participants (92% of eligible) completed the monitoring and returned their device.

### Wearable accelerometer data processing

For the baseline cohort (Cycle 11), participants were required to contribute at least four valid days of monitoring, defined as ≥21.6 hours/day (90% of the day), consistent with prior work (26,29). This threshold has been shown sufficient for stable estimation of circadian rest-activity rhythmicity from actigraphy (30,31). For the longitudinal cohort, participants were included if they had valid wear data in both Cycles 11 and 12, applying the same criteria. Using these definitions, 621 of 734 participants (83%) from the baseline cohort contributed follow-up monitoring in Cycle 12.

Raw accelerometer data from the Activity Watch were processed using the R ARCTOOLS package to summarize activity counts in minute-level epochs (32). Wear time was assessed on a day-level basis, with a valid day defined as 1,296 minutes (≥90% of the day) of wear (26). Non-wear periods were identified as ≥90 consecutive minutes of zero activity counts (26). To maximize data completeness, missing periods within otherwise valid days were imputed using the participant’s mean activity values at the same clock time across their other valid days (26).

### Assessment of circadian rest-activity rhythmicity

We derived circadian rest-activity rhythmicity biomarkers using parametric and non-parametric methods (2), providing comprehensive assessment across intensity, timing, stability, and fragmentation. These domains are consistently associated with cognition, function, and neuropsychiatric outcomes (3,4,33).

Parametric estimation employed an extended cosinor model on continuous accelerometer counts, yielding MESOR (mean activity level), amplitude (rhythm strength), acrophase (timing of peak activity), and the pseudo-F statistic (model fit to a 24-hour cycle, indicating rhythm robustness) (34).

Non-parametric indices included interdaily stability (IS; day-to-day rhythm regularity), intradaily variability (IV; rhythm fragmentation relative to a 24-hour cycle), relative amplitude (RA; normalized difference between active and rest periods), and mean activity during the most active 10-hour period (M10) and least active 5-hour period (L5) (2,35,36).

For interpretability, biomarkers were stratified into quartiles by age (≤75 vs. >75 years) and sex. For amplitude, MESOR, pseudo-F, RA, IS, and M10, higher values indicate more robust rhythmicity; therefore, Q1 (lowest quartile) was classified as non-optimal and Q2–Q4 as optimal. For IV and L5, lower values are favorable; thus, Q1 was considered optimal and Q2–Q4 non-optimal. For acrophase, earlier timing (Q1) was defined as optimal, whereas delayed timing (Q2–Q4) was considered non-optimal. Detailed cutoffs are provided in **Supplemental Table 1.** Using these definitions, longitudinal changes from Cycle 11 to Cycle 12 were categorized into four transition groups for each biomarker: Optimal (remained in the optimal range; reference), Improved (shifted from non-optimal to optimal), Deteriorated (shifted from optimal to non-optimal), and Adverse (remained in the non-optimal range).

### Assessment of functional, cognitive, and neuropsychiatric outcomes

We examined late-life functional status, cognitive performance, and neuropsychiatric outcomes. Functional status was assessed using activities of daily living (ADL; self-reported difficulty with eating, bathing, using the toilet, dressing, getting out of bed, and moving around inside the home; range 0-6, higher=greater disability) (37,38); perceived general health (“Would you say that in general, your health is excellent, very good, good, fair, or poor?”, higher=better) (39); lower-extremity function via the Short Physical Performance Battery (SPPB: gait speed, chair stand, balance; each 0-4; total 0-12, higher=better) (40); and dominant-hand grip strength (kg, maximum of two trials) (41). Cognitive performance was evaluated with immediate and delayed 10-word recall tests (each 0-10; higher=better) (29,42). To allow compatibility across outcomes, the scores were standardized to Z-scores (mean=0, standard deviation (SD)=1).

Neuropsychiatric endpoints included probable dementia and anxiety/depression. Probable dementia was defined as meeting at least one of the following criteria: (1) self- or proxy-reported dementia diagnosis; (2) cognitive impairment ≤1.5 SD below the NHATS mean in memory (≤3 on immediate/delayed recall), orientation (≤3 on naming date, month, year, day, president, and vice president), or executive function (≤1 on clock-drawing); or (3) proxy AD8 score ≥2 (43–45). For anxiety/depression symptoms, we assessed the Patient Health Questionnaire-4 (PHQ-4) and applied a cutoff ≥6, which indicates probable clinically significant symptoms, consistent with validated thresholds (29,46).

### Covariates

Covariates were selected a priori based on prior investigations linking wearable-derived activity with late-life health outcomes and were extracted from NHATS Cycle 11 (29,43). The set of covariates included age, sex (male or female), race/ethnicity (non-Hispanic White, non-Hispanic Black, other/unspecified), marital status (married/living with partner vs. not married/other), region (Northeast, Midwest, South, West), body mass index (BMI; kg/m²), daily duration of moderate-to-vigorous physical activity (MVPA; minutes/day), living arrangement (living alone vs. not), smoking status (current smoker vs. not), and comorbidities (heart attack, heart disease, or cancer).

### Statistical analysis

Survey-weighted descriptive statistics were estimated using NHATS person-level weights. Continuous variables are presented as means with standard errors (SE), and categorical variables as counts (n) with percentages (%). Variances were computed using Taylor linearization accounting for NHATS strata and primary sampling units (47).

For visualization of 24-hour circadian rest-activity rhythmicity profiles, mean minute-level activity counts were computed across the 24-hour cycle and stratified by case/control status or biomarker transition categories. Curves were smoothed with a centered rolling mean (30 minutes for baseline profiles, 90 minutes for longitudinal transitions), with shaded bands representing 95% confidence intervals. Groups were dichotomized using prespecified thresholds for functional, cognitive, and neuropsychiatric outcomes (e.g., ADL ≥2 difficulties; general health fair/poor vs. good/very good/excellent; grip strength ≤18.9 kg; SPPB ≤9; immediate/delayed recall ≤3; probable dementia present vs. absent; anxiety/depression present vs. absent). For biomarker transitions, participants were categorized as Optimal, Improved, Deteriorated, or Adverse based on changes from Cycle 11 to Cycle 12 in amplitude, MESOR, relative amplitude, pseudo-F, interdaily stability, intradaily variability, L5, and acrophase timing.

Associations between circadian rest-activity rhythmicity biomarkers and outcome measures were examined using survey-weighted generalized linear models. Incident probable dementia and anxiety/depression at one year were evaluated with survey-weighted Cox proportional hazards models, using time since Cycle 11 interview as the follow-up duration. Participants with prevalent conditions prior to Cycle 11 were excluded, and censoring occurred at death or loss to follow-up. Proportional hazards assumptions were assessed using Schoenfeld residuals (48).

All models were adjusted for age, sex, race/ethnicity, body mass index, smoking status, and baseline comorbidities (cancer, heart attack, heart disease). NHATS sampling weights were applied to generalize estimates to the U.S. older adult population. The p-values are corrected for multiple comparisons using the Benjamini-Hochberg method (49). Analyses were conducted in R (version 4.5.1) using the survey, survival, cosinor2, and nparACT packages.

## RESULTS

### Baseline characteristics

As described in **Figure 1**, a total of 747 participants contributed 7-day wearable monitoring data as part of the NHATS Cycle 11. Of these, 13 individuals were excluded due to insufficient wear time, defined as fewer than four valid monitoring days. The final analytic baseline cohort included 734 participants, contributing 5,100 person-days of data.

**Figure 1.**
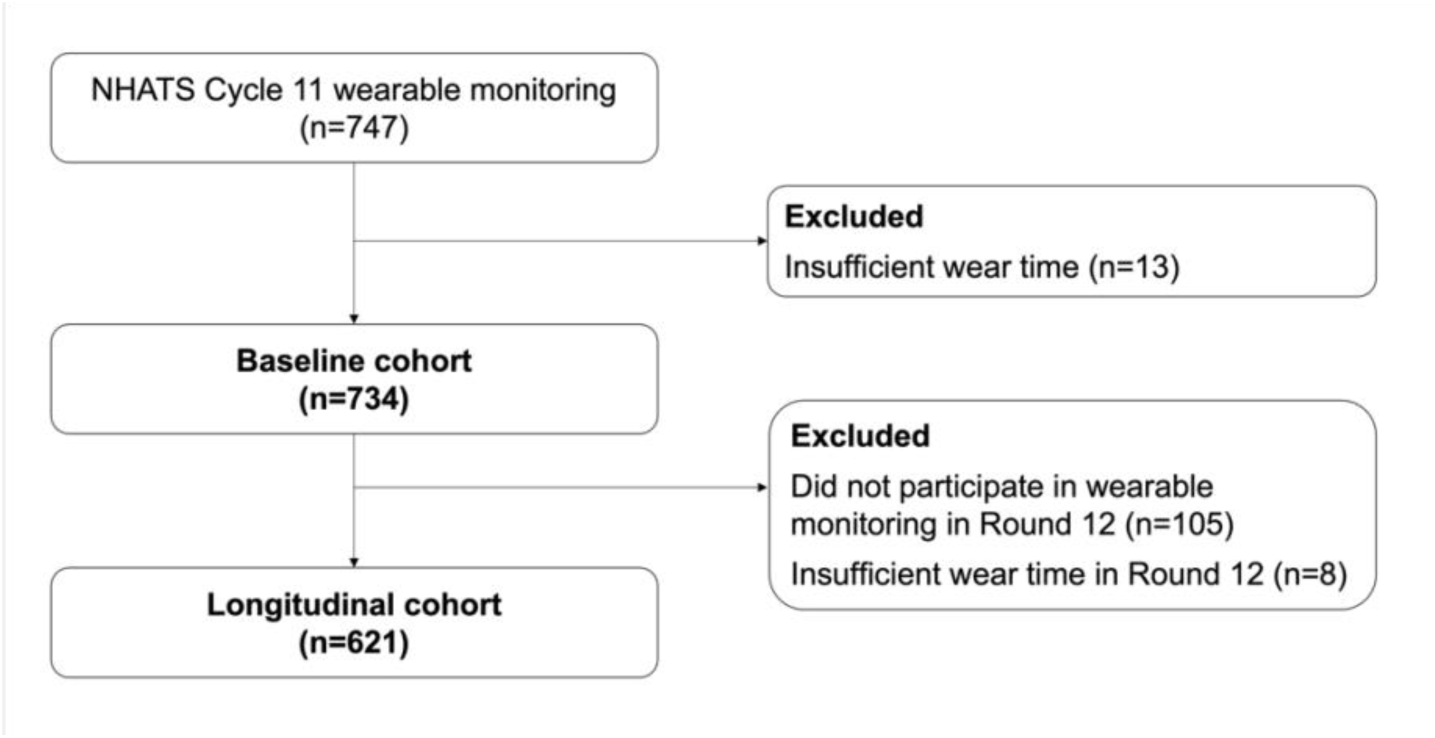
Cohort flow chart of participant selection.

To examine associations between changes in circadian rest-activity rhythmicity biomarkers and health outcomes, we further identified a longitudinal cohort that contributed valid wearable monitoring data in both Cycle 11 and Cycle 12. Of the 734 baseline participants, 105 did not participate in wearable monitoring during Cycle 12 and 8 were excluded due to insufficient wear time in Cycle 12. The final longitudinal cohort consist of 621 participants who contributed a total of 8,678 person-days of monitoring across the two cycles.

**Table 1** presents the baseline demographic, clinical, functional, cognitive, and wearable-derived digital biomarker characteristics of the cohort. The mean age was 79.2 years (SD=6.1); 55.8% were female, and 78.1% were White. Nearly half of participants were married or partnered (49.2%), and the largest proportion resided in the South (37.9%). The mean BMI was 27.9 kg/m² (SD=7.4), with participants engaging in an average of 300 minutes/day (SD=124.4) of MVPA. One-third of participants lived alone (34.4%), and most were non-smokers (94.8%). Common comorbidities included cancer (6.6%), heart attack (1.6%), and heart disease (1.6%).

**Table 1.** Baseline characteristics of the study cohort.

| Mean (SE) or N (%) | N=734 |
| --- | --- |
| **Age (years)** | 79.18 (6.11) |
| **Sex** |  |
| Female | 398 (55.8) |
| Male | 336 (44.2) |
| **Race/Ethnicity** |  |
| White | 591 (78.1) |
| Black | 57 ( 8.4) |
| Other/Not specified | 86 (13.5) |
| **Marital Status** |  |
| Married/partner | 366 (49.2) |
| Not married/other | 368 (50.8) |
| **Region** |  |
| Northeast | 131 (18.2) |
| Midwest | 167 (22.4) |
| South | 292 (37.9) |
| West | 144 (21.4) |
| **BMI (kg/m2)** | 27.94 (7.36) |
| **MVPA duration (minutes/day)** | 299.64 (124.42) |
| **Living alone** | 252 (34.4) |
| **Current smoking status** | 38 ( 5.2) |
| **Comorbidities** |  |
| Heart attack | 12 ( 1.6) |
| Heart disease | 12 ( 1.6) |
| Cancer | 51 ( 6.6) |
| **Functional status** |  |
| ADL score | 0.85 (1.59) |
| General health score | 2.71 (1.03) |
| Hand grip strength (kg) | 27.61 (12.34) |
| SPPB score | 6.95 (3.25) |
| **Cognitive performance** |  |
| Immediate word recall | 5.17 (1.68) |
| Delayed word recall | 4.06 (2.13) |
| **Possible or probable dementia** | 58 (8.2) |
| **Anxiety or depression** | 50 (7.3) |
| **Wearable digital biomarkers** |  |
| Cosinor amplitude (counts) | 1752.94 (1124.06) |
| MESOR (counts) | 1112.46 (606.76) |
| Acrophase (clock hour) | 14.42 (1.91) |
| Pseudo F (unitless) | 177.66 (154.43) |
| RA (unitless) | 0.85 (0.11) |
| M10 (counts) | 1945.11 (763.62) |
| L5 (counts) | 147.29 (147.27) |
| IV (unitless) | 0.51 (0.11) |
| IS (unitless) | 0.30 (0.08) |

Values are presented as mean (SE) for continuous variables and n (%) for categorical variables. MVPA: Moderate-to-vigorous physical activity; SPPB: Short Physical Performance Battery; RA: Relative Amplitude (unitless); M10/L5: Mean activity during the most/least active 10/5 hours; IV: Intradaily Variability; IS: Interdaily Stability. Results are weighted using NHATS survey weights.

In terms of functional status, participants had a mean ADL score of 0.85 (SD=1.59), general health score of 2.71 (SD=1.03), hand grip strength averaged 27.6 kg (SD=12.3), and a mean SPPB score of 6.95 (SD=3.25). Cognitive performance was relatively preserved, with immediate recall of 5.17 words (SD=1.68) and delayed recall of 4.06 words (SD=2.13). In Cycle 11, 8.2% (n=58) met criteria for possible or probable dementia, and 7.3% (n=50) were found positive for anxiety or depression.

Wearable-derived digital biomarkers captured circadian rest-activity rhythmicity. The mean cosinor amplitude was 1752.9 counts (SD=1124.1), MESOR was 1112.5 counts (SD=606.8), and acrophase occurred at 14.4 clock hours (SD=1.9). Rhythm robustness, reflected by the pseudo-F statistic, averaged 177.7 (SD=154.4). Nonparametric indices included relative amplitude (RA; mean=0.85, SD=0.11), mean activity during the most active 10-hour period (M10; mean=1945.1, SD=763.6), and activity during the least active 5-hour period (L5; mean=147.3, SD=147.3). Rhythm stability was assessed using intradaily variability (IV; mean=0.51, SD=0.11), indicating within-day rhythm fragmentation, and interdaily stability (IS; mean=0.30, SD=0.08), reflecting day-to-day rhythm regularity.

### Baseline 24-hour circadian rest-activity rhythmicity profiles

At baseline, participants with poorer functional and cognitive outcomes, or incident neuropsychiatric conditions, exhibited consistently lower diurnal amplitude, elevated nocturnal activity, and reduced rhythm stability, compared to their healthier counterparts (**Figure 2**).

**Figure 2.**
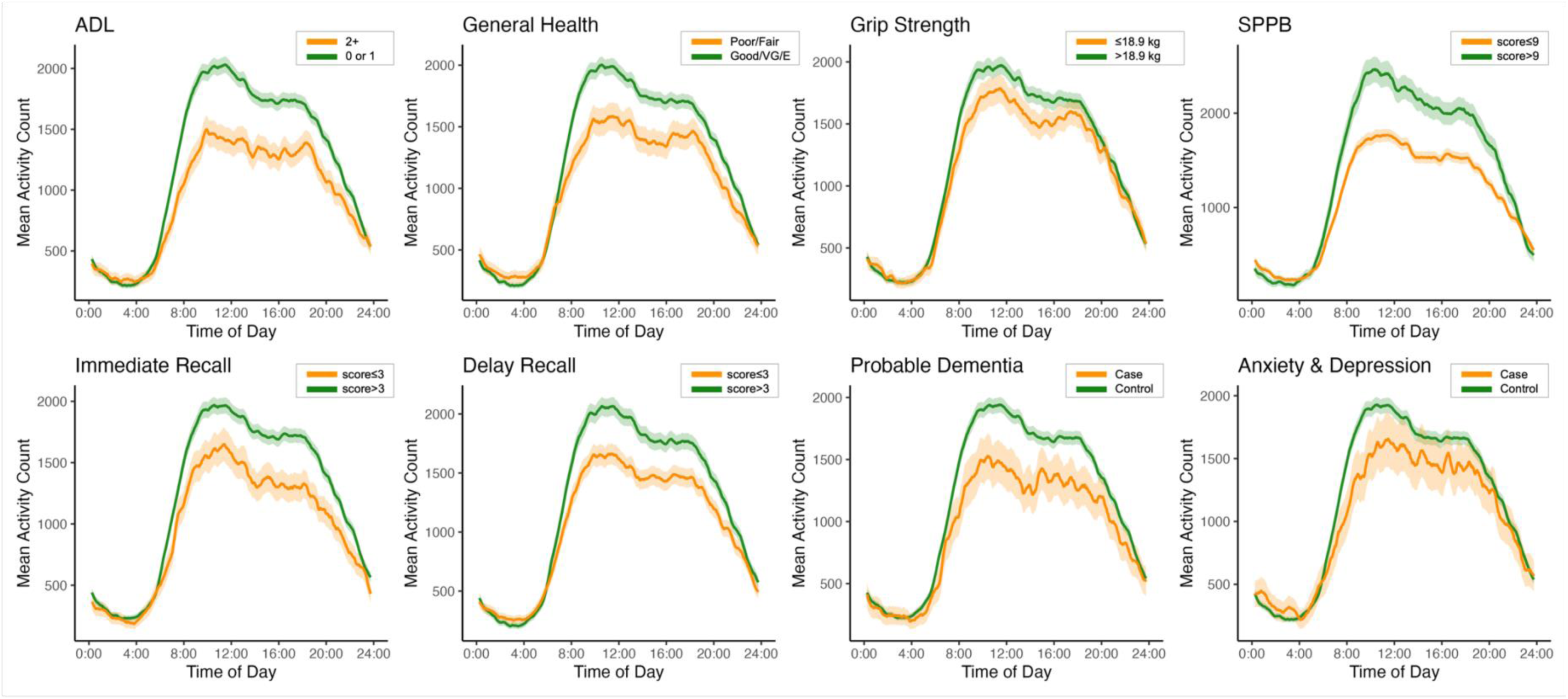
Baseline 24-hour circadian rest-activity rhythm profiles by functional status, cognitive performance, probable dementia, and anxiety/depression. Each panel displays the mean actigraphy count per minute across the 24-hour day at baseline (Cycle 11) for participants classified as cases (orange) or controls (green) for the corresponding domain (ADL, general health, hand-grip strength, SPPB, immediate recall, delayed recall, probable dementia, and anxiety/depression). Solid lines are group means; shaded bands are 95% CIs. Curves were smoothed with a centered 30-minute rolling mean. Groups were dichotomized using prespecified thresholds (e.g., ADL ≥2 difficulty; general health fair/poor vs good/very good/excellent; grip strength ≤18.9 kg; SPPB ≤9; immediate and delay recall ≤3; probable dementia present vs absent; anxiety/depression present vs absent). The x-axis shows clock time (00:00–24:00) and the y-axis shows mean activity counts.

For functional outcomes, those with ≥2 ADL difficulties or impaired physical performance (SPPB ≤9) demonstrated ∼30% lower daytime activity (1250-1350 vs. 1800-1900 counts) and markedly weaker morning peaks, indicating diminished rhythm consolidation. Participants reporting poor or fair general health also showed blunted midday activity and delayed evening decline. Lower grip strength, though less pronounced, also showed reduced daytime levels.

Cognitive vulnerability was also reflected in circadian rest-activity rhythms. Individuals with immediate or delayed recall ≤3 exhibited flattened diurnal curves, with daytime peaks of 1400 counts compared with 1800 counts in those with preserved recall.

Neuropsychiatric outcomes further reinforced these patterns. Participants who went on to develop probable dementia during follow-up had significantly lower baseline daytime activity and weaker peak amplitude, suggesting that rhythm deterioration precedes clinical onset. By contrast, those who later developed anxiety or depression showed moderately reduced daytime activity but persistently elevated nocturnal levels, with excess activity between 00:00-04:00, indicating delayed and fragmented rest-activity transitions.

Across functional, cognitive, and psychiatric domains, affected participants consistently demonstrated blunted rest-activity rhythmicity with increased rhythm fragmentation, highlighting multi-domain deterioration of circadian rhythm robustness and stability in late life.

### Baseline associations between circadian rest-activity rhythmicity biomarkers and late-life outcomes

At baseline, biomarkers reflecting rhythm intensity including cosinor amplitude, MESOR, relative amplitude, and M10 showed the most consistent associations with functional and cognitive outcomes (**Figure 3**). Participants with low amplitude reported greater ADL disability (β=0.57, 95% CI: 0.38-0.77), poorer general health (β=0.43, 0.28-0.58), weaker grip strength (β= –0.29, –0.45 to –0.14), and lower SPPB scores (β= –0.80, –0.98 to –0.62). Comparable associations were observed for low MESOR (ADL: β=0.51, 0.34-0.68; General health: β=0.41, 0.24-0.58; Grip Strength: β= –0.27, –0.41 to –0.13; SPPB: β= –0.65, –0.83 to –0.47), low RA (ADL: β=0.51, 0.26-0.76; General health: β=0.30, 0.11-0.50; Grip Strength: β= –0.34, –0.48 to –0.21; SPPB: β= –0.71, –0.87 to –0.54), and reduced M10 (ADL: β=0.56, 0.36-0.75; General health: β=0.44, 0.29-0.59; Grip Strength: β= –0.34, –0.49 to –0.19; SPPB: β= –0.73, –0.92 to –0.55).

**Figure 3.**
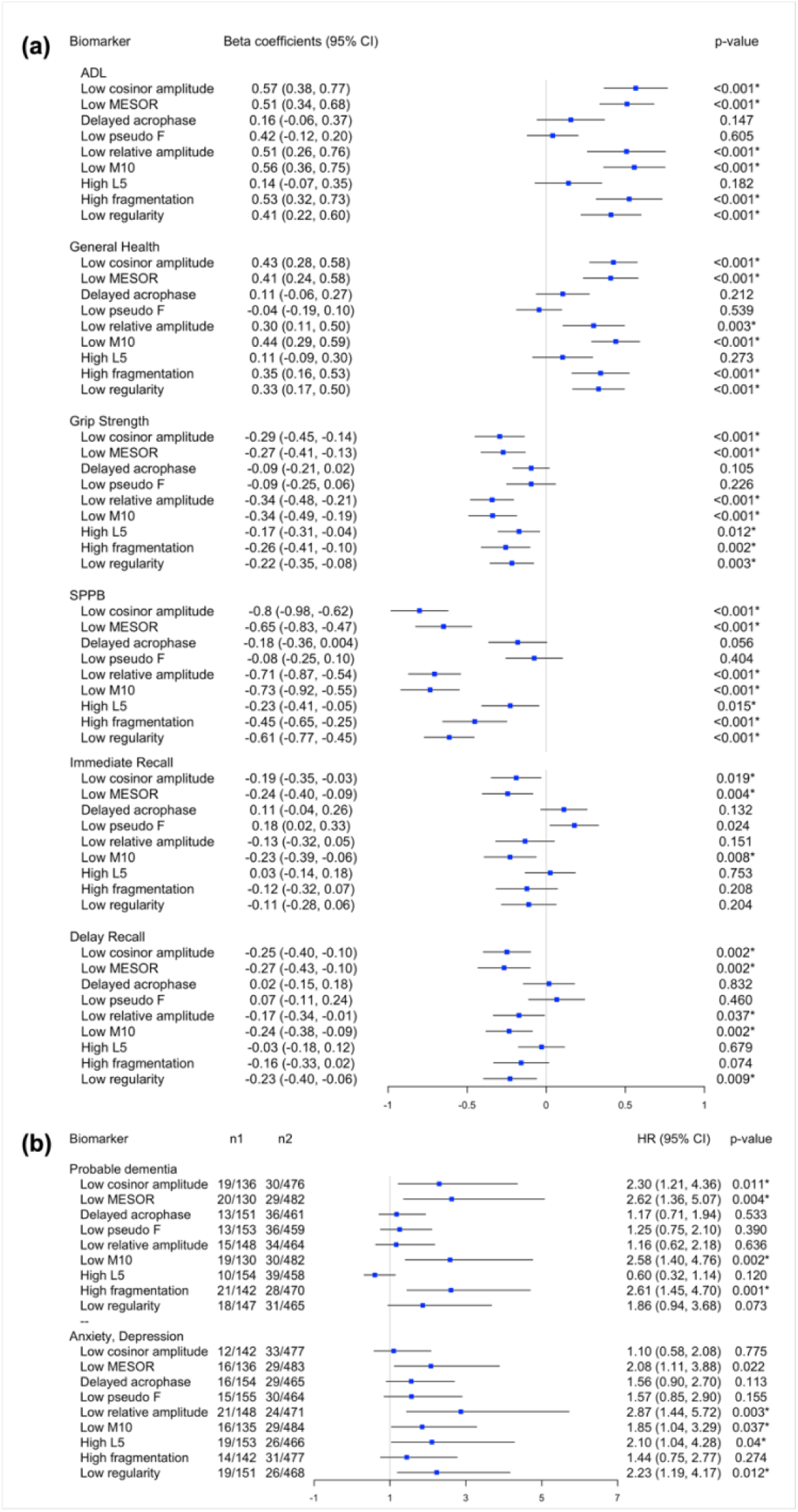
Baseline associations of circadian rest-activity rhythmicity biomarkers with late-life outcomes. **(a)** Functional status and cognitive performance; **(b)** incident probable dementia and anxiety/depression. Panel (a) presents β-coefficients with 95% CIs from survey-weighted generalized linear models, where each outcome was standardized to a Z-score. Estimates represent the adjusted mean difference (in SD units) for the “adverse” biomarker category compared with the healthier reference. For outcomes where higher values indicate worse status (ADL, general health), β>0 denotes poorer functioning. For outcomes where higher values indicate better performance (grip strength, SPPB, memory scores), β<0 denotes poorer performance. Panel (b) shows hazard ratios (95% CI) from survey-weighted Cox proportional hazards models for incident outcomes. Event counts are reported as n1/n2, where n1=number of events/total in the reference (healthier rhythmicity) group and n2=number of events/total in the adverse rhythmicity group. All models were adjusted for demographics, socioeconomics, and comorbidities and NHATS survey weights. * indicates statistical significance adjusted for multiple testing by Benjamini-Hochberg correction.

Cognition outcomes showed a similar pattern. Low amplitude, MESOR, and M10 were associated with poorer immediate recall (β= –0.19, –0.36 to –0.03; β= –0.24, –0.40 to –0.09; β= –0.23, –0.39 to –0.06, respectively) and delayed recall (β= –0.25, –0.40 to –0.10; β= –0.27; β= –0.24, –0.38 to –0.09, –0.43 to –0.10, respectively). These parallel effects suggest the central role of rhythm intensity in both physical and cognitive domains.

Beyond intensity, rhythm stability also showed robust associations across multiple domains. High IV (within-day fragmentation) was linked to greater ADL disability (β=0.53, 0.32-0.73), poorer general health (β=0.28, 0.12-0.44), lower grip strength (β= –0.26, –0.41 to –0.10), and reduced SPPB performance (β= –0.45, –0.65 to –0.25). Similarly, low IS (day-to-day regularity) was associated with higher ADL impairment (β=0.41, 0.22-0.60), poorer general health score (β=0.33, 0.17-0.50), weaker grip strength (β= –0.22, –0.35 to –0.08), poorer general health (β=0.21, 0.05-0.37), and lower SPPB scores (β= –0.61, –0.77 to – 0.45). Importantly, reduced stability also extended to cognition, with low regularity is significantly associated with a decline in delayed recall (β= –0.23, –0.40 to –0.06).

Neuropsychiatric outcomes further reinforced this distinction. For incident dementia, reductions in intensity, particularly low cosinor amplitude (HR=2.30, 1.21-4.36), low MESOR (HR=2.62, 95% CI: 1.36-5.07), and low M10 (HR=2.58, 1.40-4.76) were strong predictors, alongside high fragmentation (HR=2.61, 1.45-4.70). By contrast, risk for incident anxiety and depression was strongly associated with reduced RA (HR=2.87, 1.44-5.72), decreased M10 (HR=1.85, 1.04-3.29), and low stability (HR=2.23, 1.19-4.17).

Taken together, these results indicate that rhythm intensity markers (amplitude, MESOR, RA, M10) are dominant correlates of functional and cognitive decline, while stability measures (fragmentation, regularity) capture systemic vulnerabilities that translate into elevated risk for incident dementia and late-life anxiety or depression.

### Longitudinal changes and associations between circadian rest-activity rhythmicity biomarkers and late-life outcomes

Between NHATS Cycles 11 and 12, circadian rest-activity rhythm trajectories diverged markedly across transition groups (**Figure 4**). Participants who remained in the Optimal group preserved strong rhythms, with high daytime activity peaks and robust day-night contrast. In contrast, those who persisted in the Adverse group sustained chronically blunted profiles, with attenuated daytime activity and minimal nocturnal decline. Longitudinal changes also highlighted dynamic shifts: participants experiencing deterioration converged downward toward adverse-like profiles, while those showing improvement demonstrated partial recovery with enhanced diurnal separation. Comparable dynamics were evident for RA and pseudo-F, underscoring consistency across rhythm intensity metrics. Acrophase transitions primarily reflected timing shifts, advancing rhythms aligned with earlier peaks and higher daytime activity, whereas delayed rhythms were characterized by elevated nocturnal activity and weakened daytime peaks.

**Figure 4.**
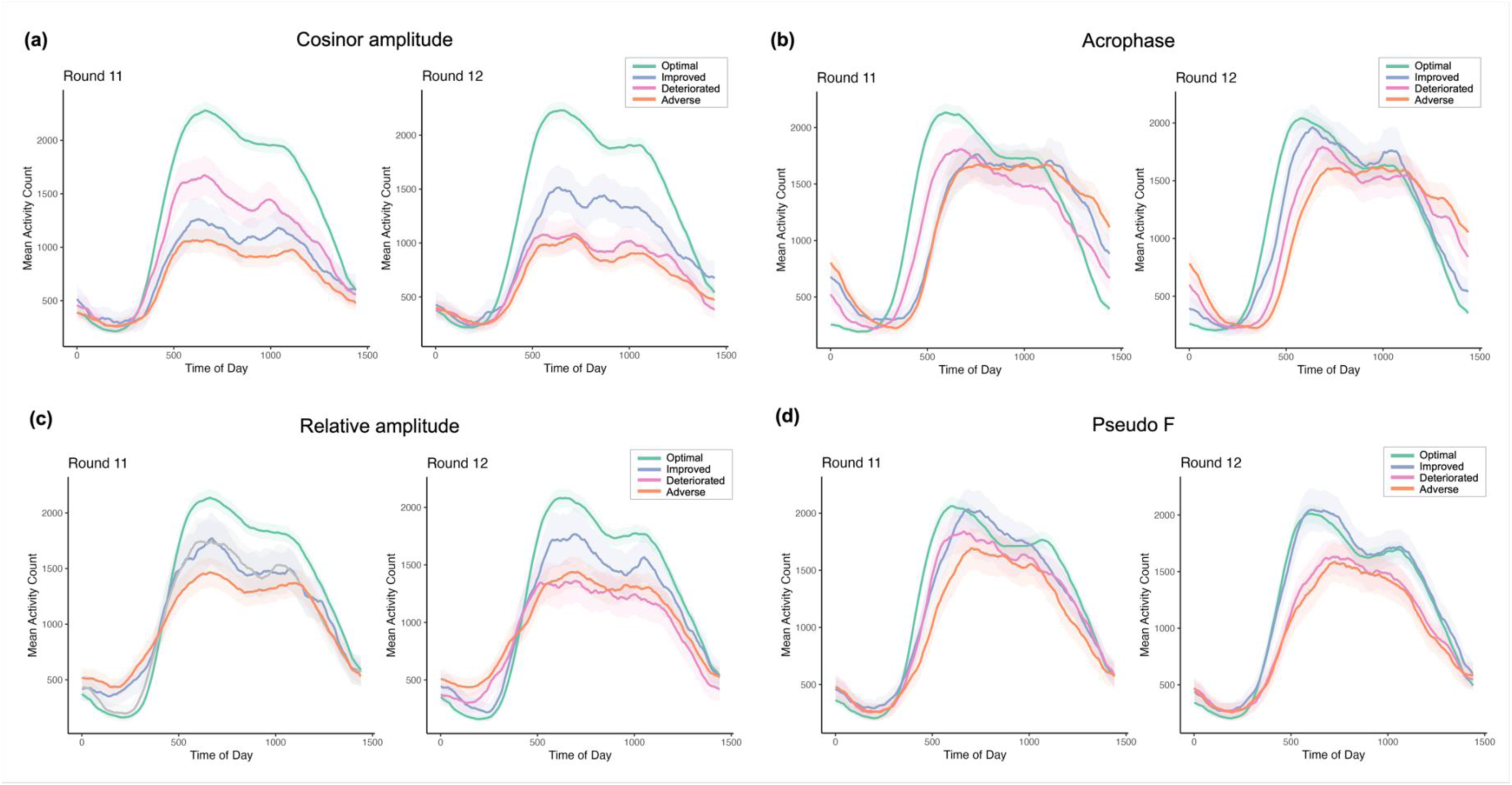
Changes in 24-hour circadian rest-activity rhythmicity profiles between NHATS Cycles 11 and 12 by biomarker transition category. Mean minute-level activity counts (solid lines) with 95% CIs (shaded ribbons) are shown for **(a)** cosinor amplitude, **(b)** acrophase, **(c)** relative amplitude, and **(d)** pseudo-F statistic. Biomarkers were stratified into age- and sex-specific quartiles (cutoffs in Supplemental Table 1) and classified as optimal vs. non-optimal: higher values (amplitude, MESOR, pseudo-F, RA, IS, M10) or lower values (IV, L5) were favorable; for acrophase, earlier timing was optimal. Transitions were defined as Optimal (optimal→optimal), Improved (non-optimal→optimal), Deteriorated (optimal→non-optimal), or Adverse (non-optimal→non-optimal). Curves were smoothed using a centered 90-minute rolling mean; the x-axis represents minutes from midnight.

In longitudinal analyses, a graded pattern of risk emerged across transition groups (**Figure 5**). Persistent adversity carried the heaviest burden, with participants in the Adverse group showing the broadest and strongest associations between circadian disruption and subsequent decline. Within this group, declines in rhythm intensity (amplitude, MESOR, M10) and stability (IS, IV) were consistently linked to worsening functional outcomes, across higher ADL disability, reduced mobility, and diminished muscle strength, as well as poorer cognitive performance, underscoring the compounding vulnerability of individuals with chronically blunted rhythms.

**Figure 5.**
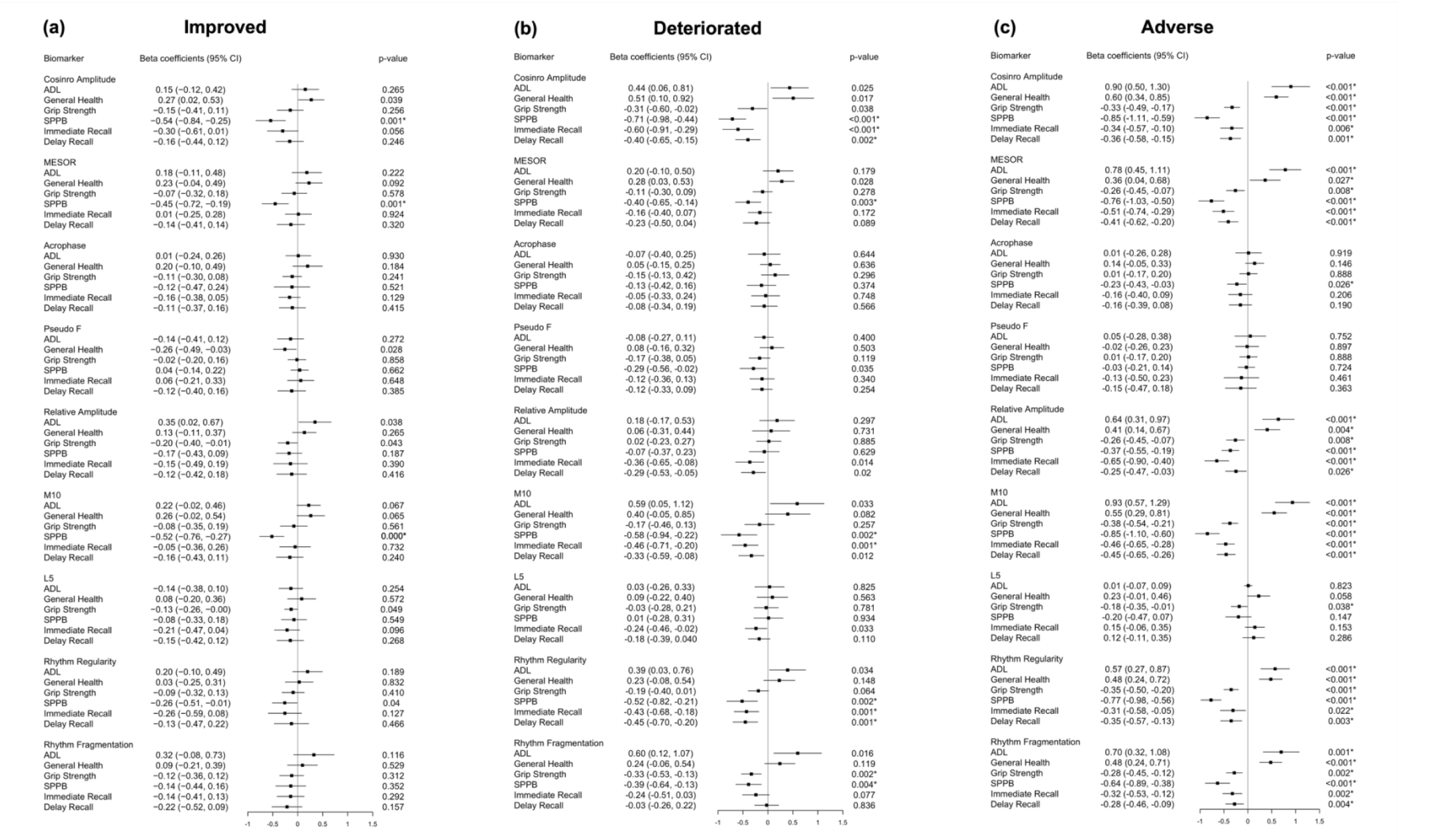
Longitudinal associations of rest-activity rhythmicity biomarkers with functional status and cognitive performance. Forest plots show β coefficients (95% CI) from multivariable models relating biomarker transitions from Cycle 11 to 12. Outcomes were standardized (z-scores); β>0 denotes worse ADL/health, whereas β < 0 indicates poorer grip strength, SPPB, or memory. All models were adjusted for demographics, socioeconomics, and comorbidities and NHATS survey weights. * indicates statistical significance adjusted for multiple testing by Benjamini-Hochberg correction.

Deterioration represented the most dynamic risk state. Participants who shifted toward adverse-like profiles exhibited marked losses across both functional and cognitive domains. Declines in rhythm intensity, particularly cosinor amplitude, were strongly linked to adverse late-life outcomes, including lower SPPB performance (β= –0.71, –0.98 to –0.44) and diminished immediate recall (β= –0.60, –0.91 to –0.29) even after adjusting for covariates and multiple testing. Similar patterns were observed for MESOR and M10 deterioration that reductions significantly indicated poorer SPPB performance and immediate recall.

Stability markers reinforced these findings. Declining rhythm regularity are strongly associated with decreased functional (SPPB: β= –0.52, –0.82 to –0.21) and impaired cognition, affecting both immediate (β= –0.43, –0.68 to –0.18) and delayed recall (β= –0.45, –0.70 to –0.20). Increased rhythm fragmentation uniquely captured adverse effects on functional outcomes, including reduced grip strength (β= –0.33, –0.53 to –0.13) as well as reduced SPPB (β= –0.39, –0.64 to –0.13). In contrast, deterioration in acrophase, pseudo-F, RA and L5 displayed weaker and insignificant associations after multiple testing correction.

Improved transitions generally moved in the opposite direction, pointing toward better outcomes. Although associations seldom reached statistical significance, improvements consistently aligned with stabilization of function and cognition, suggesting that even partial recovery may mitigate the trajectory of decline.

Finally, the significance map confirmed this hierarchy (**Supplemental Figure 1**). Adverse transitions exerted the broadest and strongest associations, with nearly all biomarkers, except acrophase, pseudo-F, and L5, predicting functional and cognitive decline. Deteriorated transitions carried robust risks, particularly in rhythm intensity and stability indices. Improved transitions indicated more favorable directions. Taken together, these findings underscore that rhythm worsening, whether persistent or progressive, remains the reliable driver of late-life functional and cognitive decline, while stabilization or partial recovery may help offset further losses.

## DISCUSSION

In this large, nationally representative cohort of older adults, we demonstrated that circadian rest-activity rhythmicity is a robust predictor of late-life health trajectories. The main findings are threefold. First, weaker rhythm intensity and stability at baseline were consistently associated with functional impairment, cognitive deficits, and elevated risk of neuropsychiatric outcomes. Second, longitudinal analyses revealed a graded hierarchy of risk. Participants with persistently non-optimal rhythms experienced the most severe deterioration, while those who worsened converged toward similarly adverse profiles. By contrast, individuals with improving rhythms did not worsen and, across several domains, showed signals of stabilization, underscoring that change in rhythmicity, rather than static levels alone, most strongly inform late-life vulnerability. Third, rhythm domains contributed both jointly and differentially: low intensity was the core determinant across outcomes, while high fragmentation was more strongly linked to probable dementia, and poor day-to-day stability was more closely tied to depression and anxiety.

Our findings extend prior research in several important ways. Accumulating evidence has demonstrated that reduced circadian amplitude and poor stability are associated with adverse outcomes, including mortality (50), frailty (51), dementia (52,53), cognitive decline (54), and depression (55) in general population. Among older adults, the central role of amplitude has been particularly emphasized. In the Baltimore Longitudinal Study of Aging, lower amplitude was significantly associated with faster memory decline in a cohort of 422 adults aged ≥50 years (56). A smaller study of 43 cognitively intact older adults reported that higher daily activity was associated with better memory and executive functioning (57). Consistent with these findings, a larger population-based analysis of 2,317 middle-aged and elderly participants found that major depressive disorder was characterized by significantly weaker rhythmicity, particularly lower amplitude (58).

Despite their importance, prior investigations were largely cross-sectional, typically based on a single week of actigraphy. While informative, such designs cannot capture the dynamic alterations in circadian rest-activity rhythmicity that signal early vulnerability to physiological dysregulation and late-life health deterioration. By incorporating repeated circadian assessments in a nationally representative cohort, our study provides rigorous evidence that within-person deterioration in rhythm intensity and stability is significantly associated with functional and cognitive decline. Equally important, the observation that improvement conveyed stabilization indicates that circadian disruption is not an inevitable feature of aging but a potentially modifiable process.

The plausibility of these associations is supported by converging biological pathways. Blunted amplitude, for example, reflects weakened physiological oscillations and loss of multi-system synchrony, both hallmarks of biological aging (2). This weakening is often accompanied by greater fragmentation, which signals instability in circadian control, and by reduced day-to-day stability, which reflects impaired coupling between central and peripheral clocks (3,7). Circadian disruption also fuels chronic low-grade inflammation, a process strongly implicated in dementia, depression, and frailty (17,59–61). These vulnerabilities are further compounded by common exposures in late life, including insufficient physical activity, prolonged sedentary behavior, and diminished sensitivity to light, all of which erode circadian regulation (1,2). Altogether, these mechanisms offer a coherent biological explanation for why circadian deterioration accelerates functional and cognitive decline and emphasize that interventions targeting these pathways may enhance resilience, preserve late-life performance, and ultimately delay phenotypes of morbidity and mortality.

Our study carries important translational implications. First, circadian rest-activity rhythmicity can be passively and continuously captured through wearables and smartphones, positioning digital phenotyping as a scalable and accessible tool for monitoring aging populations in real-world settings. Second, the integration of circadian measures with high-resolution activity data provides a rich substrate for advanced computational and statistical methods, including machine learning and deep learning, which can facilitate individualized risk stratification and the design of adaptive, personalized interventions. Third, participants whose rhythms deteriorated exhibited the most pronounced losses across functional and cognitive domains, whereas those with improving rhythms showed relative stabilization. This pattern indicates that circadian rest-activity rhythmicity is not merely a static marker of vulnerability but a dynamic and potentially modifiable target for intervention. Building on this, enhancing rhythm intensity through structured physical activity and morning light exposure, and improving stability through consistent daily routines and sleep-wake scheduling, represent practical strategies to preserve cognitive and functional capacity in older adults. These insights position circadian rest-activity rhythmicity as a digital biomarker for early risk detection and as an actionable target to promote resilience and extend healthspan in late life.

The strengths of this study include its use of repeated circadian measures in a large, nationally representative cohort of U.S. older adults and its linkage of both baseline levels and longitudinal changes to diverse late-life outcomes ranging from ADL and grip strength to dementia and depression. This multidimensional design enhances generalizability and provides robust evidence for causal inference. Nonetheless, several limitations warrant consideration. The two-year follow-up window may not capture long-term trajectories, though exclusion of individuals with possible dementia or depression at baseline helped mitigate reverse causality. The number of incident dementia and psychiatric cases was modest, reflecting the smaller subset with wearable monitoring. Furthermore, actigraphy provides indirect measures of circadian rest-activity rhythmicity and did not include detailed sleep parameters. Future studies with longer follow-up, larger samples, and multimodal circadian assessments are warranted.

In conclusion, our findings establish circadian rest-activity rhythmicity as dynamic determinants of late-life outcomes. Persistent weakness or deterioration of rhythmicity predicted accelerated decline in function and cognition, whereas improvement conveyed stabilization. Leveraging repeated circadian assessments and digital phenotyping in a large, nationally representative cohort, we demonstrate that these biomarkers serve not only as early indicators of vulnerability but also as actionable targets for intervention. Embedding circadian digital biomarkers into clinical and population research provides a scalable, accessible, and noninvasive framework for early detection and precision interventions, with the potential to transform aging research into dynamic, real-world monitoring that sustains independence, cognitive resilience, and healthy longevity.

## ABBREVIATIONS

ADL: Activities of Daily Living
BLSA: Baltimore Longitudinal Study of Aging
BMI: Body Mass Index
CI: Confidence Interval
HR: Hazard Ratio
IS: Interdaily Stability
IV: Intradaily Variability
L5: Mean activity during the least active 5-hour period
M10: Mean activity during the most active 10-hour period
MESOR: Midline Estimating Statistic of Rhythm
MDD: Major Depressive Disorder
MVPA: Moderate-to-Vigorous Physical Activity
NHATS: National Health and Aging Trends Study
PHQ-4: Patient Health Questionnaire-4
RA: Relative Amplitude
SD: Standard Deviation
SE: Standard Error
SPPB: Short Physical Performance Battery

## AUTHOR CONTRIBUTIONS

J.S. contributed to conceptualization, methodology, data curation, formal analysis, writing (original draft), writing (review and editing). J.P.O. contributed to methodology, data interpretation, writing (review and editing).

## DATA AVAILABILITY

The dataset supporting the conclusions of this article are available at https://nhats.org/researcher/data-access. The codes are available upon request to the corresponding author.

## DECLARATIONS

### Ethics approval and consent to participate

NHATS was approved by the Johns Hopkins Bloomberg School of Public Health Institutional Review Board (IRB No. 00002083). All NHATS participants completed an informed consent. The data used for this research were de-identified and publicly available.

### Consent to participate

Not applicable

### Completing Interests

The authors declare no competing interests.

